# Social, functional and quality-of-life outcomes among long term acute care hospital survivors with tracheostomy

**DOI:** 10.64898/2026.01.20.26343699

**Authors:** Hiam Naiditch, Amanda Moale, S. Mehdi Nouraie, Bryan J. McVerry, Snigdha Jain, Anna Zemke

## Abstract

**Rationale:** Patients receiving prolonged mechanical ventilation are often discharged to long-term acute care hospitals (LTACHs) with hopes of recovery and ultimately return to the community. Among those who survive and undergo tracheostomy, little is known about their quality of life and social outcomes after LTACH discharge.

**Objective:** Measure health related quality of life in a cohort of critical illness survivors who underwent tracheostomy and an LTACH stay

**Methods:** Single center, prospective observational cohort study conducted at a long-term acute care hospital between 2022-2024. Adults with prolonged mechanical ventilation requiring tracheostomy were eligible. Survivors or surrogates completed a telephone survey 3-6 months after LTACH admission. Surveys included the Katz Index of Independence in Activities of Daily Living (ADL) and Patient-Reported Outcome Measurement Information System (PROMIS) measures of physical, mental, and social health. Descriptive statistics summarized scores; exploratory analyses examined associations between persistent tracheostomy and residence.

**Results:** Seventy participants were enrolled (median age 64 years; 58.6% male). Median ICU length of stay was 40 days prior to LTACH transfer. By LTACH discharge, 39 people (56%) had been decannulated. At a median follow-up of 5.5 months, 50 participants (71%) were alive, 39 completed the survey. Most respondents reported impairments in feeding, dressing, and bathing. PROMIS t-scores demonstrated severe impairments in physical function (median 26.7), and ability to participate in social roles (37.0), with high symptom burden of depression (61.9) and anxiety (58.3). Greater impairment was observed among participants with ongoing tracheostomy or were not residing at home. Home residence did not differ significantly by decannulation status (61.8% decannulated vs. 46.7% not decannulated; OR 1.85 (95% CI 0.54, 6.30)).

**Conclusions:** Survivors of PMV and LTACH admission experience marked functional impairments, restricted participation in social roles, and prominent symptoms of depression and anxiety—particularly among patients not living at home or with persistent tracheostomy. These data may help clinicians prepare patients and families for life after tracheostomy and guide tailored support addressing multifaceted needs.

**Primary Funding Source:** National Institutes of Health

## Introduction

Approximately 4 million patients who survive acute critical illness each year, of whom 5-10% go on to develop chronic critical illness (CCI)(1). A common feature of chronic critical illness (CCI) is prolonged mechanical ventilation (PMV) with placement of a tracheostomy tube to facilitate long-term mechanical ventilation. Patients requiring tracheostomy and PMV require intensive rehabilitation and exhibit markedly higher odds of transfer to a long-term acute care hospital (LTACH) for ventilator weaning and rehabilitation (2) (3)(4).

Despite 30 years of health care advances, nearly half of all patients requiring tracheostomy for PMV die within 1 year(5)(6)(7)(8). Among ICU survivors discharged to LTACHs, impaired physical functioning is common; nearly half remain dependent for activities of daily living (ADLs) two years following LTACH admission(9). Outcomes are particularly poor for those who undergo PMV requiring tracheostomy, experiencing 0-3 median days alive and out of an institution by day 90 from ICU admission, indicating a high reliance on healthcare facilities(10). PMV is also associated with severe impairments in self-perceived peripheral strength and objectively measured ability to perform ADLs which are predominant in the first 6 months following LTACH discharge (11). Health-related quality of life is often similarly impaired. Mirroring post-intensive care syndrome (PICS), nearly half of all individuals admitted to an LTACH report symptoms of anxiety and depression, with many patients additionally experiencing impaired cognition, post-traumatic stress disorder, and overall lower HRQoL(12) (13)(14). Yet few studies have adequately captured patient-reported social outcomes such as social participation and satisfaction in conjunction with functional and other HRQoL outcomes, prompting the Society for Critical Care Medicine consensus statement identifying this as an area needing more research (15). Moreoever—while social outcomes have been previously explored among all ICU survivors using such tools as EQ-5D—the experience of patients discharged to an LTACH who require tracheostomy for PMV remains understudied(16)(17)(18). Furthermore, whether removal of a tracheostomy tube (“decannulation”) is associated with LTACH survivor outcomes is unknown.

To better characterize patient experiences following PMV, we examined patient-reported social, HRQoL, and functional outcomes among LTACH survivors requiring tracheostomy for PMV. Further, we explored how outcomes differ by tracheostomy decannulation status by LTACH discharge and disposition at 3-6 months. Understanding social outcomes and their correlates among patients undergoing PMV considering tracheostomy is key to providing anticipatory guidance for patients, families, and caregivers, while providing a basis for strenghthening social connectedness among LTACH survivors(18)(18,19)(20). Moreover, identifying which outcomes are associated with persistent tracheostomy or LTACH survivor disposition informs tailoring of interventions by patient subgroup across care settings.

## Methods

### Study design

Data were collected as part of a single-center prospective cohort study at an LTACH in Pennsylvania evaluating tracheobronchitis host-pathogen biology in parallel with patient-participant and surrogate surveys of physical, social and mental health. The study was approved by the University of Pittsburgh IRB (STUDY 20110443); informed consent was obtained from participants or surrogates when participants were incapacitated per study protocol (see Supplement). Survey data was deidentified at collection and stored securely. This work presents the patient-reported physical, social, and mental health outcomes at 3-6 months following LTACH admission and follows the Consensus-based Checklist for Reporting of Survey Studies (CROSS)(21).

### Data collection methods

As a part of the parent study (see Supplement), we collected baseline demographics, pre-admission functional status, and social support, and conducted a follow-up survey 3-6 months post-enrollment. Clinical data including decannulation status at LTACH discharge were extracted from the electronic medical record (**Table 1**); additional clinical and socioeconomic characteristics are included in the Supplement (**Table S1**).

### Post discharge survey

Physical, mental, and social functioning were assessed using the Katz Index of Independence in Activities of Daily Living (Katz ADL) and Patient-Reported Outcome Measurement Information System (PROMIS) instruments, both of which have been used to evaluate outcomes among ICU survivors (**Supplemental Methods**) (22) (23)(24)(25)(26) (27). The Katz Index rates functional independence across six basic self-care tasks—bathing, dressing, going to toilet, transferring, continence, and feeding—with binary responses (yes/no) for independence in each category; total score 0-6 (6: independent; 4: moderate impairment; ≤2: severe impairment(28). The PROMIS scales assess an array of patient-reported functional and quality of life outcomes. Guided by existing literature and our study team’s expertise, the following PROMIS measures were selected for our survey: Sleep Disturbance v1.0, Pain Interference v1.1, Fatigue v1.0, Cognitive Function-Abilities Subset v2.0, Physical Function v2.0, Ability to Participate in Social Roles and Activities v2.0, Emotional Distress – Anxiety v1.0, and Emotional Distress Depression v1.0(29)(30)(31)(32)(33)(34)(35). The PROMIS Physical Function scale was included to assess a broader range of functional impairment as a complementary measure to Katz, which primarily identifies severe impairment. PROMIS t-scores are normalized to a population mean of 50.

### Sampling and administration

We recruited adults aged >=18 years who were admitted to the LTACH with a tracheostomy between August 2022 and December 2024. Individuals with a diagnosis of cystic fibrosis or history of lung transplantation were excluded, as these represent distinct patient populations for the purposes of the parent tracheobronchitis study. Baseline surveys were administered in person by a trained research coordinator during LTACH admission. Post-LTACH follow-up surveys were administered via telephone by a research coordinator between 3-6 months after enrollment. PROMIS measures were administered using computer adaptive testing (CAT). Surveys were completed by patient-participants when possible or by or with the help of their surrogate decision-maker.

### Statistical Analysis

Descriptive statistics were used to summarize baseline characteristics and outcomes. Katz ADL and PROMIS t-scores are reported as medians and interquartile ranges (IQR). PROMIS scores were compared to the reference population mean of 50 using two-sided z-tests. Discharge disposition among survivors was dichotomized as home vs. all other dispositions due to sample size. Patient-level transitions across disposition categories over time were visualized using alluvial diagrams. Subgroup analyses contrasted decannulated vs. persistent tracheostomy at discharge and home vs. other residence at follow-up; Fisher’s Exact Test was used due to sample size. The association between decannulation at discharge residence at home was assessed using the Pearson correlation coefficient, and unadjusted odds ratios were obtained via logistic regression. Correlograms were constructed to display pairwise correlations among PROMIS domains. Additional bivariate analyses evaluated the association between number of people a patient could discuss important matters prior to critical illness and PROMIS social functioning t-score. Descriptive statistics were calculated using Stata version 18.0 (College Station, TX). GraphPad PRISM version 10 (Boston, MA) and R packages *ggplot2* (version 3.4.0) and *ggalluvial* (version 0.12.5) were used for analyses.

### Missing Data

Complete case analysis was chosen to evaluate correlations in our exploratory analyses. The limitations of the statistical approach are noted in the Discussion.

## Results

### Cohort description

A total of 70 participants (median age 64 years, male (58.6%)) were enrolled over a period of 28 months.(**Table 1**; **Figure S1)**. Medical comorbidities are described in **Table 1**, and socioeconomic factors are reported in **Table S1.** Among eligible participants, 29% consented and were enrolled a median of 5 days after LTACH admission [IQR 4, 11] and 89 days [IQR 68, 136] after index hospital admission. Tracheostomy was typically performed by acute hospital day 18 [IQR 11, 22 days]. Median LTACH LOS was 41 days [IQR 26, 62]. 60 participants (85.7%) completed the baseline survey. 39 participants (76% of survivors) completed the follow-up survey (**Figure S1**).

At the time of the first/baseline study visit (29/69), 41% of participants were fully liberated from the ventilator but had a tracheostomy in place. By LTACH discharge, 39 (56%) were decannulated; 10 (14.5%) had non-respiratory barriers to decannulation (e.g. subglottic stenosis or high cervical spinal cord injury). At follow-up (phone survey 5.5 months from LTACH admission [IQR 4.7, 6.5 months]), 27% of participants had died. Survivors had a significantly shorter ICU LOS than non-survivors (median 35 vs 65 days, p=0.004). Among survivors, n=26 (52%) were first discharged to inpatient rehabilitation (IPR). At follow-up, n=28 (56%) lived at home (**Figure 1**). Participants decannulated at discharge were more likely to be discharged to IPR (65.7% vs. 18.8% not decannulated; OR 7.94 (95% CI 1.73, 51.92; p = 0.003)) but were not more likely to reside at home (60.0% vs. 43.8%; OR 1.90 (95% CI 0.50, 7.62)). Decannulation status did not show a statistically significant correlation with discharge home (Fisher’s exact p=0.53)).

**Figure 1:**
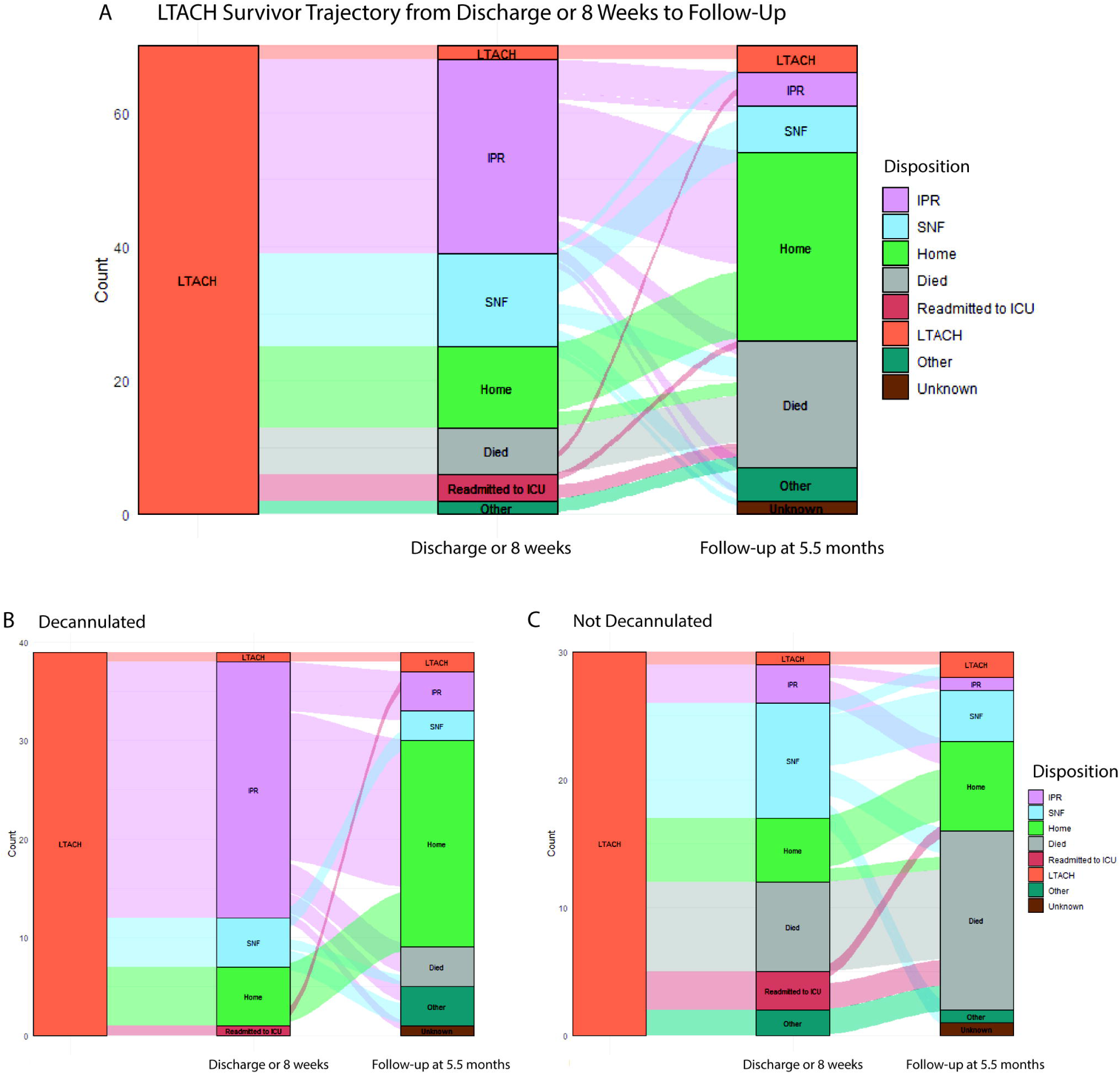
The location of residence was determined at 8-weeks (or LTACH discharge if that occurred prior to 8 weeks), and at a median of 5.5 months from study enrollment in all participants (A) as shown in the alluvial plot. Disposition by tracheostomy cannulation status is shown in (B) and (C).

### Profound functional impairment is seen in LTACH survivors at 6 months

Prior to critical illness, 73% (44/60) of the cohort were fully independent in all ADLs (Katz Index 6, **Figure 2A**); 94% lived at home. At follow-up, survivors commonly reported impairments in feeding, continence, dressing, and bathing (**Figure 2A**). PROMIS scores for functional impairment were markedly reduced (median 26.7 [IQR 19.1, 41.3])) (**Figure S3D**). Participants discharged from the LTACH with a tracheostomy, regardless of subsequent decannulation, had worse functional impairment as measured by Katz (**Figure 2B**) and PROMIS (**Figure 2D**). Home residence at follow-up was associated with greater ADL-independence (**Figure 2C**) and improved physical function T-scores (median 36.4 [IQR 26.1, 43.5] vs. 20.2 [IQR 15.2, 24.3]) (**Figure 2D**).

**Figure 2:**
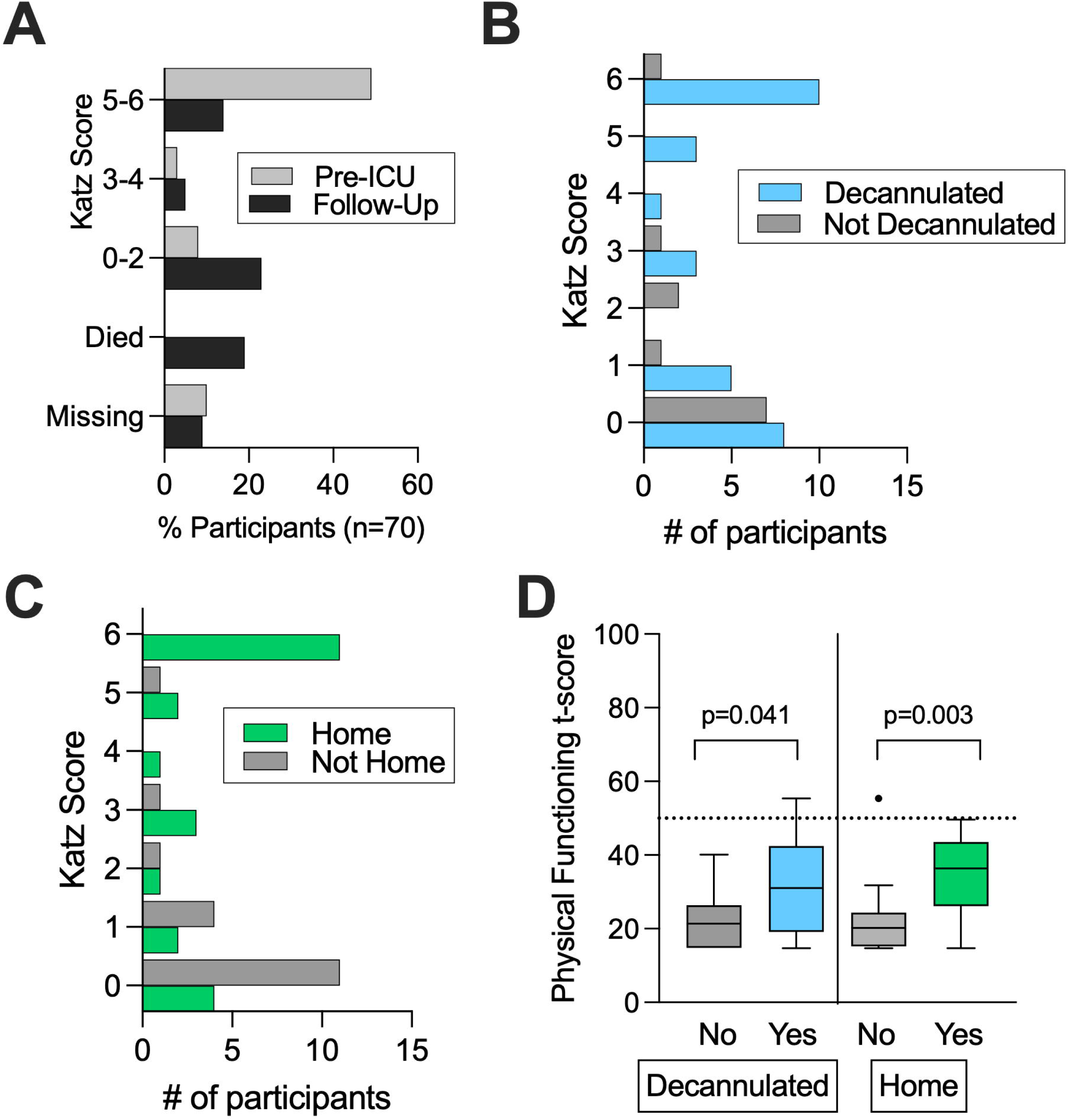
(A) Katz Activities of Daily Living (ADL) scores from prior to ICU admission and upon follow-up. (B) Katz scores shown by decannulation status at LTACH discharge. (C) Katz scores shown by residence at home at follow-up phone call. (D) PROMIS Physical Functioning t-scores shown by decannulation status at LTACH discharge and by residence at home on follow-up phone call survey. P-values reported from Mann-Whitney test.

### The ability to participate in social roles and activities is impaired following discharge

Social functioning, measured by the PROMIS Ability to Participate in Social Roles and Activities scale, was markedly reduced among survivors (median T-score 37.0 [IQR 26.1-45.8]; **Figure S3C**). Persistent tracheostomy showed a non-significant association with lower social functioning (p=0.085) (**Figure 3A**). Survivors residing at home showed significantly higher social functioning as compared to those not residing at home (median 43.8 [IQR 37.0, 54.0] vs 26.5 [IQR 21.5-30.5], p<0.0001) (**Figure 3B**). Prior to illness, participants reported a median of 2 ([IQR 1, 3], maximum 10) people to talk to about important matters. Having more than one person with whom to discuss important matters prior to illness showed a non-significant association with improved social functioning after discharge (p=0.062) (**Figure 3B**). Physical functioning and social participation were strongly correlated (Spearman coefficient=0.90; **Figure 4**).

**Figure 3:**
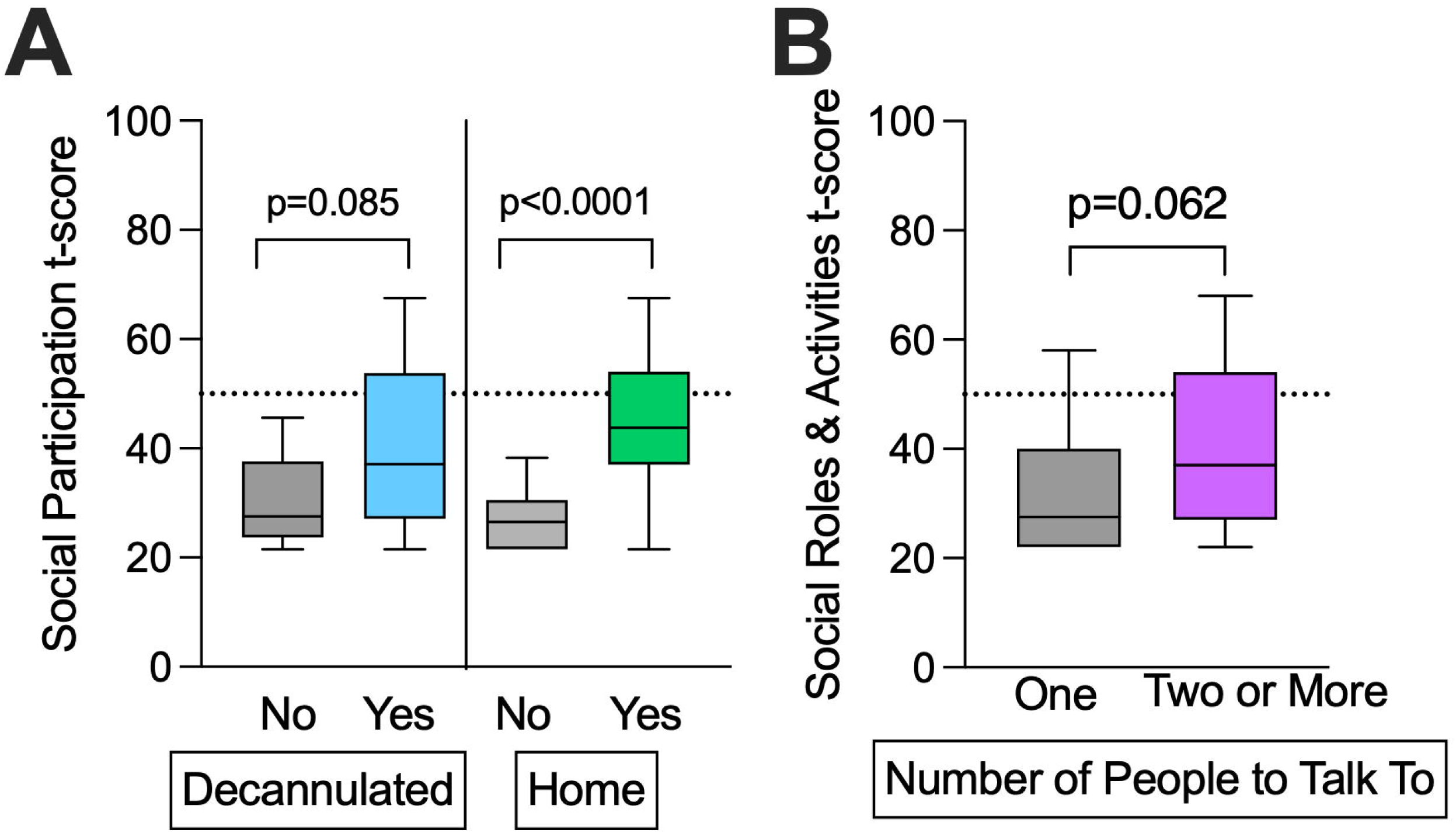
(A) PROMIS Social Functioning t-scores shown by decannulation status at LTACH discharge and by residence status at follow-up. (B) PROMIS Social Functioning t-scores at follow-up shown by number of people to talk to about important matters prior to hospitalization for critical illness. P-values from Mann-Whitney test.

**Figure 4.**
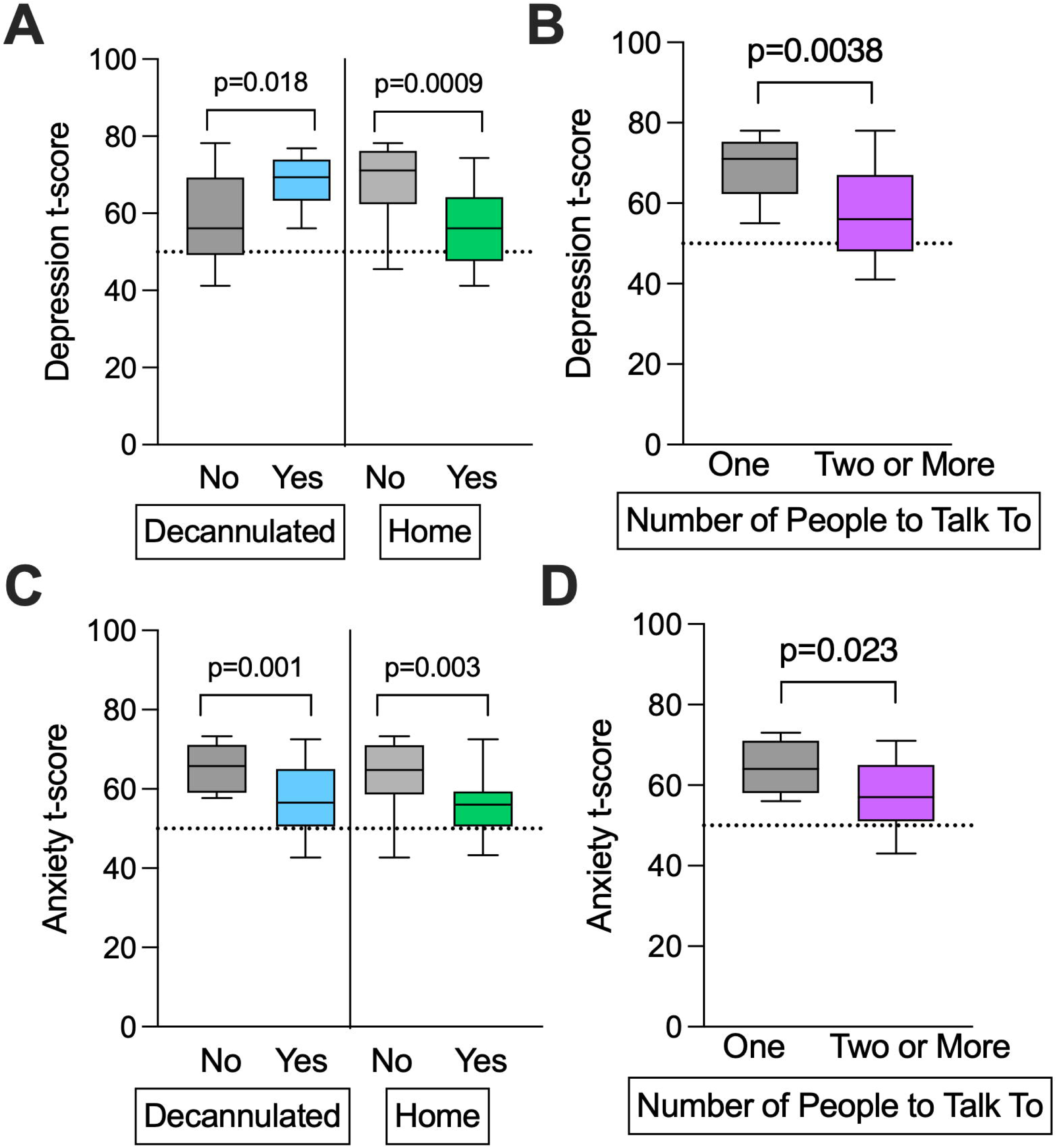
Plot of correlation matrix for depression, anxiety, physical functioning, social functioning. and other quality of life symptoms among LTACH patients who underwent tracheostomy. Spearman correlation coefficients shown within each box.

Tracheostomy and inability to be discharged home were associated with adverse psychiatric and HRQoL outcomes Among survivors, PROMIS T-scores indicated higher depression (**Figure S3A**) and anxiety (**Figure S3B**) compared to population norms. Higher symptom burden was observed among participants not decannulated at LTACH discharge (**Figure 5A, 5C**), those not living at home at follow-up (**Figure 5A, 5C**), and those who had only a single person with whom they could discuss important matters (**Figure 5B, 5F**). Among other PROMIS-reported HRQoL symptoms, pain (p=0.028) and fatigue (p=0.015) were significantly elevated compared to population norms. Cognitive Function-Abilities scores was lower (T-score 44.7; p=0.001). Sleep disturbance did not differ from norms (p=0.480)(**Figure S4**).

**Figure 5:**
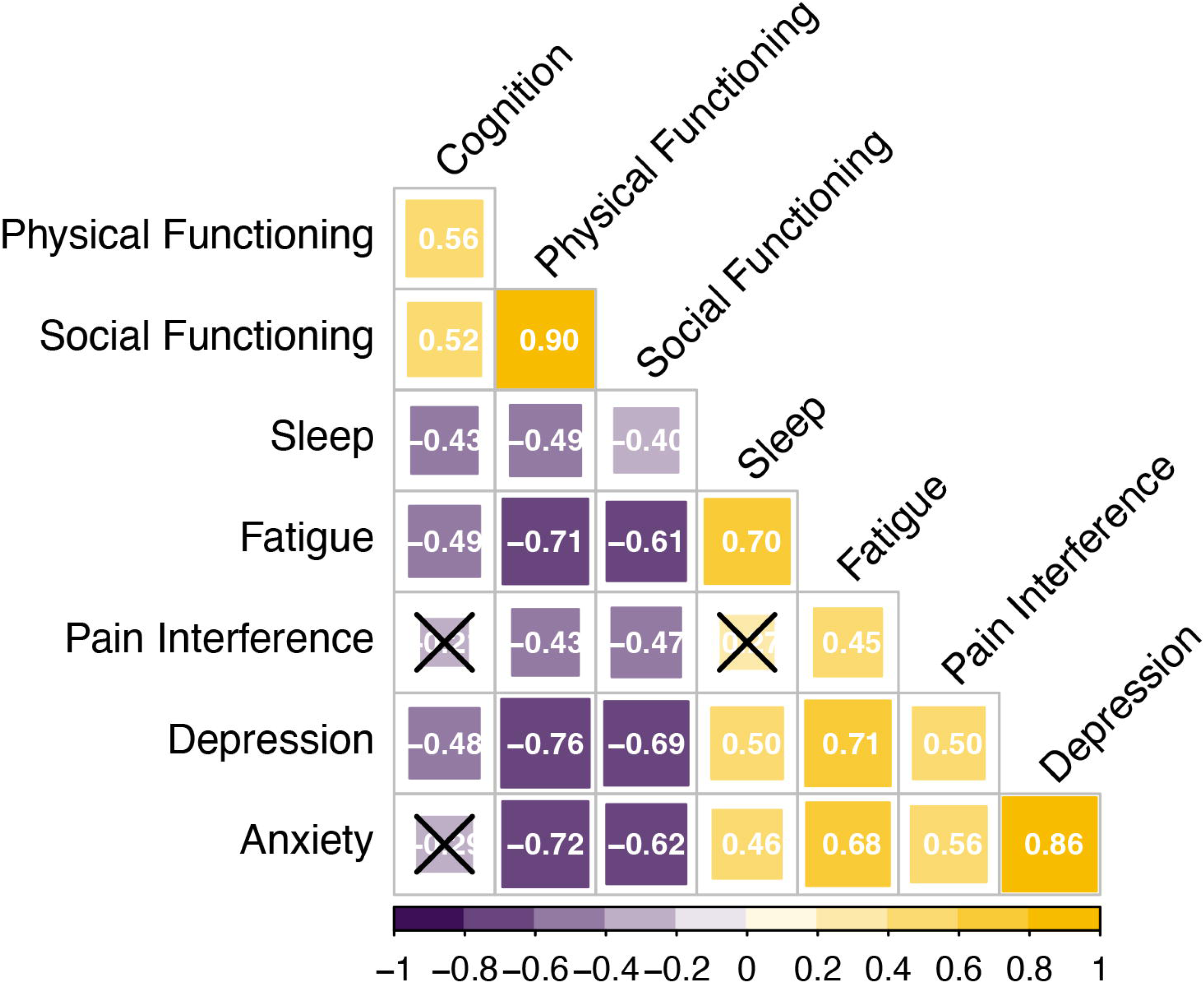
(A) PROMIS Depression t-scores shown by decannulation status at LTACH discharge and by residence status at follow-up. (B) PROMIS Depression t-scores at follow-up shown by number of people to talk to about important matters prior to hospitalization for critical illness. (C) PROMIS Anxiety t-scores shown by decannulation status at LTACH discharge and by residence status at follow-up. (D) PROMIS Anxiety t-scores at follow-up shown by number of people to talk to about important matters prior to hospitalization for critical illness. The expected value of the reference population (50) is shown as a dotted red line for reference. P-values from Mann-Whitney test.

## Discussion

In a single-center cohort study of PMV survivors with tracheostomy surveyed 3-6 months after LTACH admission, we observed a significant burden of depression, anxiety, impaired physical functioning, and limited ability to participate in social roles relative to the general population. Not living at home on follow-up was associated with impaired physical functioning, functional independence, social functioning, depression, anxiety, pain and fatigue. Persistent tracheostomy at LTACH discharge was associated with impaired physical function, greater ADL dependence, depression, and anxiety, but not impaired social functioning. Our study builds on previous research by elucidating functional, social and mental health outcomes in LTACH survivors who undergo tracheostomy after PMV, a population less well-studied. Consistent with international society recommendations that emphasize social functioning as a core component of PICS, we describe novel findings on social functioning among survivors of tracheostomy and PMV(15). Collectively, these findings can inform expectations for patients and their caregivers while supporting clinicians in planning tailored interventions to promote recovery and improve their quality of life.

LTACH patients with tracheostomy show substantial deficits in functional, mental health, and other HRQoL domains, which parallel sequelae reported after critical illness. Consistent with previously reported physical impairment in 15-50% of ICU survivors at 12 months, physical functioning in our cohort up to six months from LTACH admission remained severely impaired (9,11)(36). Similarly, we found a high prevalence of anxiety in our cohort, aligning with reports showing symptoms 6-12 months post-discharge among one third of critical illness survivors (37)(38)(39)(40). The high rate of moderate-severe depression in our cohort may reflect higher initial acuity, prolonged medical course, or the ongoing presence of a tracheostomy tube(41). The low cognitive scores in our cohort may be influenced by inclusion of participants with severe neurologic insult (e.g., anoxic brain injury) as their index illness. Similarly, pain and fatigue were also elevated, consistent with prior post-ICU literature(42) (43).

Importantly, we found that social functioning is impaired among LTACH survivors, particularly among those with persistent tracheostomy. While prior studies included social health as a subdomain, such studies do not adequately isolate the unique and relatively common subset of patients who experience PMV, tracheostomy and discharge to LTACH(44). Moreover, the use of PROMIS allows for a more granular assessment of social participation and satisfaction—constructs not fully captured by EQ-5D or SF-36—potentially explaining why some studies reported no clear healthcare-related impact on social functioning among LTACH survivors who undergo PMV(14)(45)(46)(47). We additionally build on qualitative work by Eakin et al emphasizing the relevance of PROMIS domains among survivors acute respiratory distress syndrome ARDS and acute respiratory failure (26). Social functioning is a key component of PICS, and can impact quality of life, mental health, and community reintegration(15). Social isolation has been shown to be a risk factor for adverse outcomes such as increased mortality, increased disability burden, and placement in a skilled nursing facility(18). Importantly, participants who reported having only a single person with whom they could discuss important matters prior to their illness reported significantly greater symptoms of depression, underscoring the role of perceived social support and aligning with SCCM statements that the “absence of social support across the illness is a key risk factor for mental health problems post-ICU”(15). To improve social functioning and support, in line with previous work by Falvey et al among geriatric survivors of critical illness(18), we propose routine screening for social isolation and participation and dysfunction among LTACH patients, and advocate for more robust studies exploring the impact of peer support interventions including in-person or virtual support groups, “buddy” programs, and volunteer phone calls aimed to increase social connectedness both inside the LTACH and upon discharge(18)(18,19)(20). Implementaiton would likely be best guided by an interdisciplinary model engaging social workers and rehabilitation specialists in providing comprehensive care for survivors of critical illness as a whole (20,48). Indeed, health care facilities such as hospitals have been previously recognized as ideal sites at which social isolation can be identified and addressed through individual and community-level interventions including support groups and peer-to-peer volunteer involvement (18). Given that tracheostomy is associated with a substantially fewer days alive outside of an institution, facility-level strategies to mitigate social isolation may be especially important for this population (10).

The strong correlation between physical functioning and the ability to participate in social roles and activities, although unsurprising, suggests a role for social support systems in promoting return to functional and mental health recovery as seen in other health conditions like stroke, trauma, and chronic disease. Conversely, functional impairment may inhibit access to medical, psychiatric and rehabilitation services while also impacting an individual patient’s ability to socialize. Although causal directions are not possible to untangle in our observational study, this finding warrants further exploration to elucidate recovery mechanisms among LTACH survivors and to test multifaceted interventions that target both physical and social recovery.

Our findings have practical implications for patients, caregivers, and clinicians. Patients residing at home at follow-up and those decannulated at LTACH discharge reported improved physical function and social participation and lower levels of anxiety and depression. These data can inform expectations early in a patient’s hospital course and inform discharge and follow-up planning at LTACHs. Decision-making surrounding tracheostomy placement among ICU survivors can be complex, with limited discussion regarding long-term outcomes(50).

Understanding expectations can help support survivors of critical illness and their families through what is often a complex recovery process, and help support providers through goal setting and informed anticipatory guidance(12). Furthermore, identifying sequelae of critical illness can guide interventions in the community for LTACH survivors. Our data indicate that management of mood disorders following discharge is a particular need, and the suggest that care fragmentation that occurs during transitions out of the ICU may further complicate mental health care for PMV survivors(7). Reinforcing screening to systematically identify symptoms of depression and anxiety among PMV patients, particularly those geared towards managing symptoms across care transitions, might improve mental health outcomes. Moreover, given the role social isolation may play in modifying patient outcomes, our work supports a role for studies of novel interventions that enable survivors to participate in social activities within the context of evolving physical limitations. Future studies might also elucidate patient-centered perspectives on social participation and other quality of life outcomes to optimize such interventions(10).

### Limitations

Our findings should be interpreted in the context of several limitations. First, the small sample size (n=70) and single-center design may limit generalizability. Second, in the absence of instruments validated specifically among LTACH survivors, we used well-established measures of physical, mental, and social functioning among adults and ICU survivors overall. These instruments would benefit from continued validation among LTACH survivors who undergo PMV, particularly as we noted unanticipated potential floor and ceiling effects within the PROMIS scales. Third, although use of telephone surveys has been validated (51), this mode may lead to selection bias toward patients with adequate access or cognitive and communication ability to use telephone communication. Fourth, a mix of participants and their surrogates completing the phone surveys due to practical considerations. Fifth, while our initial protocol planned for surveys to be conducted at 3 months (120 days) following LTACH admission, logistical constraints—similar to those reported by Gardner et al (49)—shifted most follow-up to 5-6 months. Finally, we acknowledge that our findings should be viewed as exploratory given the use of complete case analysis and the potential for survivorship bias, and further acknowledge—as noted by Law and Walkey—that those patients who did not survive an LTACH or PMV may have substantially different experiences and quality-of-life considerations that fail to be adequately captured in a study such as ours (50).

### Conclusion

In a cohort of LTACH survivors following tracheostomy tube placement and PMV, a high burden of limitations in physical function, depression, and anxiety and a reduced HRQoL and ability to participate in social roles were identified. Persistent tracheostomy and non-home residence were associated with worsened outcomes. These results can help clinicians guide expectations of patients and families and identify resources for supporting recovery at LTACH discharge.

## Data Availability

All data produced in the present work are contained in the manuscript

## Acknowledgements

We thank the patients and families who generously volunteered their time and energy while walking down very difficult paths. We also thank the clinical staff at the study site for their support. Delana Walters provided invaluable assistance as study coordinator.

## Disclosures

ACZ, BJM, HN, and AM have received funding from NIH. BJM has received funding from Genentech unrelated to this work. ChatGPT was used to assist with coding in R, for selective sentence structuring, and to assist with adherence to word count requirements.

## Abbreviations

PROMIS: Patient-Reported Outcome Measurement Information System
ADL: Activities of daily living
LTACH: Long term acute care hospital
HRQoL: health-related quality of life
LOS: length of stay
PMV: Prolonged Mechanical Ventilation
PICS: Post Intensive Care Syndrome

## Notes

### Funding Statement

This study was funding by the National Institutes of Health

### Author Declarations

University of Pittsburgh Institutional Review Board gave ethical approval for this work.

