## Supplementary material for "Social, functional and quality-of-life outcomes among long term acute care hospital survivors with tracheostomy": Table 1

**Table 1. Key cohort characteristics**

| **Age, years (median [IQR])** | 64.50 [57.00, 69.75] |
| --- | --- |
| **Sex at birth, n (% male)**  Female  Male | (n=70)  29 (41.4)  41 (58.6) |
| **Race, n (%)**  White  Black  Asian | (n=70)  60 (85.7)  9 (12.9)  1 (1.4) |
| **Insurance, n (%)**  Medicare  Medicaid  Private  Unknown | (n=60)  33 (55.0)  14 (23.3)  12 (20.0)  1 (1.7) |
| **Pre-admission residence, n (%)**  Home  Skilled nursing facility | (n=66)  62 (94.0)  4 (6.06) |
| **Lived alone prior to admission, n (%)**  No  Yes | (n=64)  47 (73.4)  17 (26.6) |
| **Initial Cause of Respiratory Failure, n (%)**  Medical Illness  Post-Surgical  Neurologic Illness | (n=70)  34 (48.6)  22 (31.4)  14 (20.0) |
| **Medical Comorbidities, n (%)**  *Neurologic*  Severe Neurologic Injury  Dementia  Developmental Disorder  Depression  Spinal Cord Injury  Bipolar disorder or Schizophrenia  Neuromuscular disease  Stroke  *Cardiopulmonary*  Chronic Obstructive Pulmonary Disease  Obesity Hypoventilation Syndrome  Prior home positive pressure device  Prior home oxygen  Atrial fibrillation  Coronary artery disease  Congestive Heart Failure  CPR prior to LTACH admission  *Renal and Endocrine*  End Stage Kidney Disease on Dialysis  Chronic kidney disease (not on dialysis)  Diabetes mellitus  *Recent Surgical History*  Open heart surgery  Abdominal or thoracic surgery  Severe trauma requiring surgery  Vascular surgery  *Other*  Malignancy (excluding skin cancers)  Remote liver or kidney transplant  Known contraindication to tracheostomy decannulation | (n=70)  8 (11.4)  2 (2.9)  3 (4.3)  19 (27.1)  7 (10.0)  2 (2.9)  8 (11.4)  11 (15.7)  12 (17.1)  4 (5.7)  8 (11.4)  2 (2.9)  25 (35.7)  22 (31.4)  17 (24.3)  21 (30.0)    17 (24.3)  6 (8.6)  25 (35.7)  14 (20.0)  14 (20.0)  3 (4.3)  3 (4.3)    26 (37.1)  6 (8.6)  10 (14.5) |
| **ICU length of stay, days (median [IQR])** | 40.00 [26.00, 60.00] |
| **Decannulated by LTACH Discharge, n (%)** | 39 (56.5%) |
| **Relationship of Surrogate to Participant, n (%)**  Spouse/partner  Parent  Child  Sibling  No surrogate participated in the study | (n=70)  26 (37.1)  7 (10)  8 (11.4)  7 (10)  22 (31.4) |

No participants identified as Hispanic or Latino. Self-identified sex at birth and race are reported. The variables living alone prior to admission, insurance status, residence prior to admission were obtained solely from participant/surrogate interview and had missing data as indicated
