## Supplemental Information for "Social, functional and quality-of-life outcomes among long term acute care hospital survivors with tracheostomy"

**Social, functional and quality-of-life outcomes among long term acute care hospital survivors with tracheostomy: A single center prospective cohort study**

Supplementary Index

**Table of Contents:**

**Page number. Content**

**2.**	**Supplemental Methods**

**5.**	**Figure S1.** Modified Consort Diagram

**6.**	**Table S1.** Socioeconomic Description

**8.**	**Figure S2:** Timeline of participants’ flow through medical system

**9.** **Figure S3:** PROMIS depression, anxiety, physical functioning and ability to participate on social roles and activities results.

**10. Figure S4:** PROMIS sleep, fatigue and pain scale results

**Supplemental Methods**

**Study Description**

The manuscript reports an observational cohort study conducted in a 30-bed hospital-within-hospital long term acute care hospital (LTACH) in Pennsylvania. The study was IRB-approved by University of Pittsburgh (STUDY20110443). Inclusion criteria were having a tracheostomy present on admission, ability to obtain consent, and age >18 years. Exclusion criteria were a diagnosis of cystic fibrosis or lung transplant because these disease conditions are known to modify the airway microbiome from prior research. Our university requires the treating medical team to obtain permission for study personnel to approach before any discussions around potential study participation, which required close coordination and buy-in from the on-site, interdisciplinary medical staff. The study site census was screened through weekly contact with staff that had usual clinical access to the medical record. Consent for study participation was obtained by trained study personnel, either the research coordinator or a study physician. Because of the high rate of delirium and other cognitive impairments in chronic critical illness, surrogate consent was obtained if an individual was not determined to have decisional capacity through discussions between the research staff and treating medical team. If a participant later regained capacity, assent to continue participation was obtained. We also offered study participation to the participant’s support person (surrogate-decision maker, legally authorized representative) and consented these individuals separately for participation in joint interviews around social determinants of health and health related quality of life measures.

After study enrollment, a weekly visit was done where a sterile suction kit was used to obtain an undiluted tracheal aspirate and blood samples were banked. Study visits occurred for a maximum of 8 weeks. For participants still admitted to the LTACH at 8 weeks from consent, their discharge location at 8 weeks was listed as “LTACH”. The medical history was extracted using a standardized form from the medical record on both admission and discharge because new medical problems were frequently identified during the LTACH stay and admission paperwork could be incomplete. Admission type (medical, surgical, neurologic) was adjudicated by medical record review. During admission at the LTACH and participants or their support person completed a questionnaire around pre-ICU functional status and social determinants of health that was administered in person by the research coordinator. These data were available for 60/70 (86.7%) of the cohort, with missing data resulting from very early discharges or transfers back to the ICU. Participants were able to decline to answer any particular question. At a median of 122 days from LTACH admission, the PROMIS measure questionnaires were completed over the telephone with the research coordinator. We also determined the disposition of the participant at that point. Deaths were determined through the medical record covering our region and internet death notice or obituary searches.

**Description of Outcome Measures:**

The Katz Index assesses an individual’s ability to perform six basic self-care tasks: bathing, dressing, going to toilet, transferring, continence, and feeding. Responses are binary (yes/no) for independence in each category, with a total score of 6 indicating full function, 4 indicating moderate impairment, and 2 or less indicating severe impairment. The PROMIS scale is a battery of updated measures of patient-reported outcomes including both functional and quality of life outcomes such as self-reported pain, emotional distress, and ability to participate in social roles. PROMIS scales are reported as t-scores relative to the population mean of 50, with higher values dependent on the context of the measure (e.g. a higher depression score indicates worse depression, while a higher physical functioning score indicates better physical functioning). The following tools were used: Sleep Disturbance version 1.0, Pain Interference version 1.1, Ability to Participate in Social Roles and Activities version 2.0, Emotional Distress – Anxiety version 1.0, Emotional Distress Depression version 1.0, Fatigue version 1.0, Cognitive Function – Abilities Subset version 2.0, Physical Function version 2.0. Computerized adaptive testing was use for these scales, which adjusts the question order to achieve the most precise results in the fewest questions. All measures used had been previously designed for CAT implementation. Data were recorded in a custom RedCap database hosted by the University of Pittsburgh.

Software Used: Descriptive statistics were calculated using Stata version 18.0 (College Station, TX). Data were visualized using GraphPad PRISM version 10 (Boston, MA) and the R packages *ggplot2* (version 3.4.0) and *ggalluvial* (version 0.12.5).

**Figure S1:**

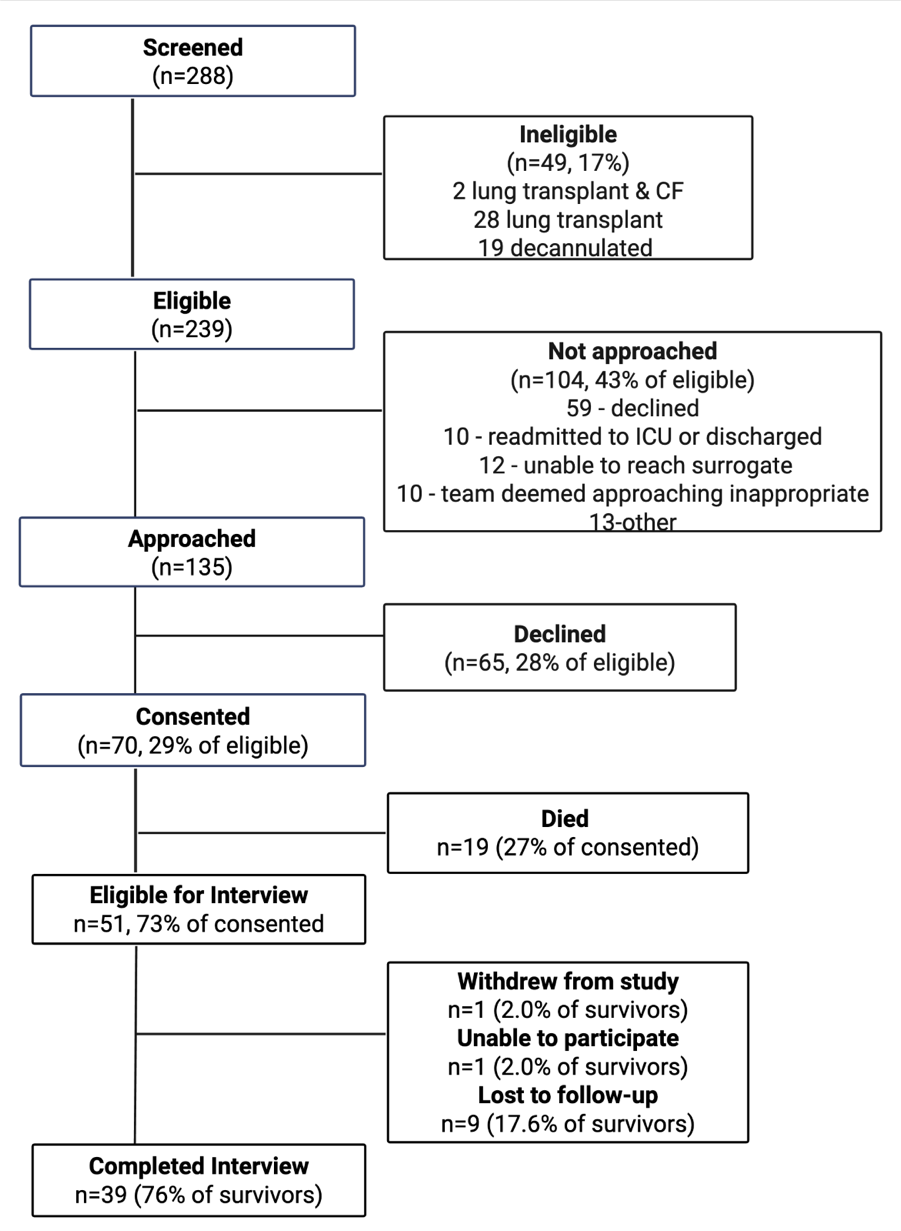

**Figure S1:** Flow chart of screening, enrollment, and survey participation data for this study.

**Table S1:** Additional socioeconomic description of cohort

|  | N (%) |
| --- | --- |
| How confident are you filling out medical forms by yourself?  Extremely  Quite a bit  Somewhat  A little bit  Not at all | (N=60)  29 (48.3)  14 (23.3)  5 (8.3)  8 (13.3)  4 (6.7) |
| How many people did the participant have to talk about important things?  1  2  3 or more | (N=60)  16 (26.7)  21(35.0)  23 (38.3) |
| Before this illness, how many minutes away was the primary caregiver?  0  5-20  120 | (N=59)  46 (78.0)  12 (20.0)  1 (1.69) |
| How far away (in minutes) is the support person from the LTACH  10-25  30-60  61-120  More than 2 hours | (N=58)  22 (37.9)  27 (46.6)  4 (6.9)  5 (8.6) |
| At the end of the month, does the participant’s household have:  More than enough money left over  Just enough | (N=60)  11(18.3)  49 (81.7) |
| Do you have FMLA available (for the caregiver)?  No  Yes  Unsure | (N=60)  53 (88.3)  4 (6.7)  3 (5.0) |
| Employment status before ICU admission  Employed  Retired  Unable to work  Unemployed | (N=63)  19 (30.2)  27 (42.9)  14 (22.2)  3 (4.8) |
| Who completed the PROMIS surveys?  Participant  Surrogate  Both  Missing (data was not being collected yet) | (N=39)  13 (33.3)  7 (17.9)  10 (25.6)  9 (23.1) |

Participants and their caregivers rarely reported problems with transportation for medical appointments (1.7%), social activities (1.7%), or generally (3.4%). All participants had electricity and water. Most participants had broadband internet (93.3%). All participants spoke English as their primary language. Abbreviations: *FMLA: Family and Medical Leave Act; ICU: Intensive Care Unit; PROMIS: Patient-Reported Outcomes Measurement Information System.*

**Figure S****2 : Summary of Time Course for participants by survival status**
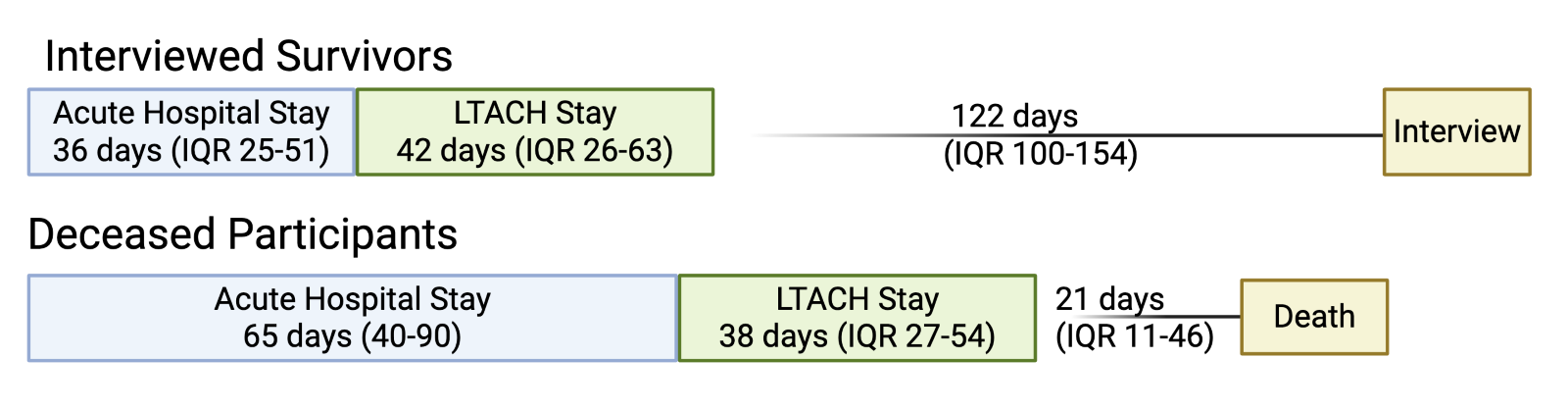

**Figure S2: Summary of Time Course for participants by survival status.** Participants who survived until the interview timepoint had significantly shorter acute hospital (ie ICU) stays as compared to those who died prior to the interview.

**Figure S3: PROMIS score histograms**

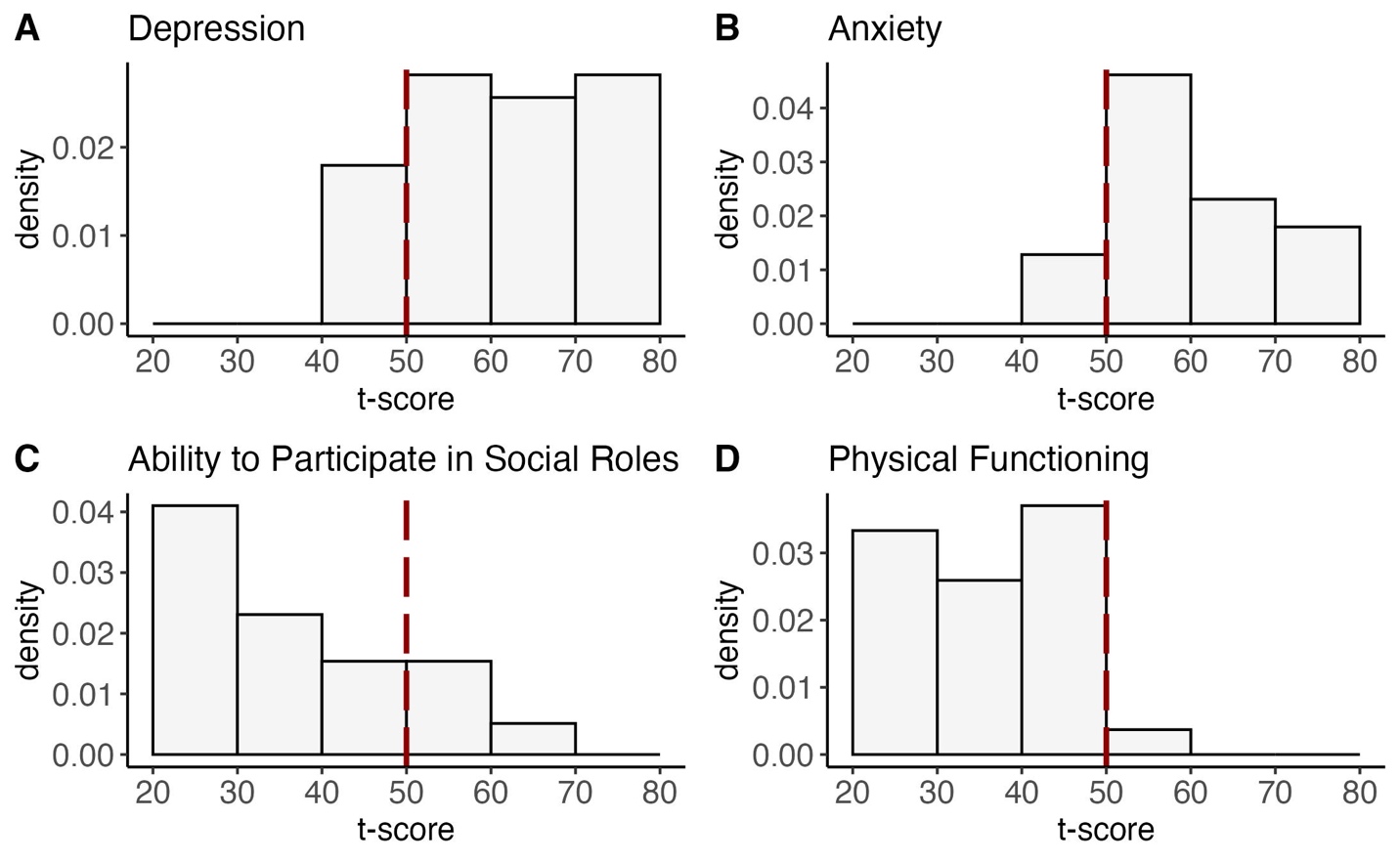

**Figure S3: PROMIS score histograms**. Depression (A), Anxiety (B), Ability to Participate in Social Roles and Activities (C), and Physical Functioning (D) scales are as shown. The reference population is plotted in red as a normal distribution with a mean of 50 and standard deviation of 10.

**Figure S4: Additional PROMIS score histograms**

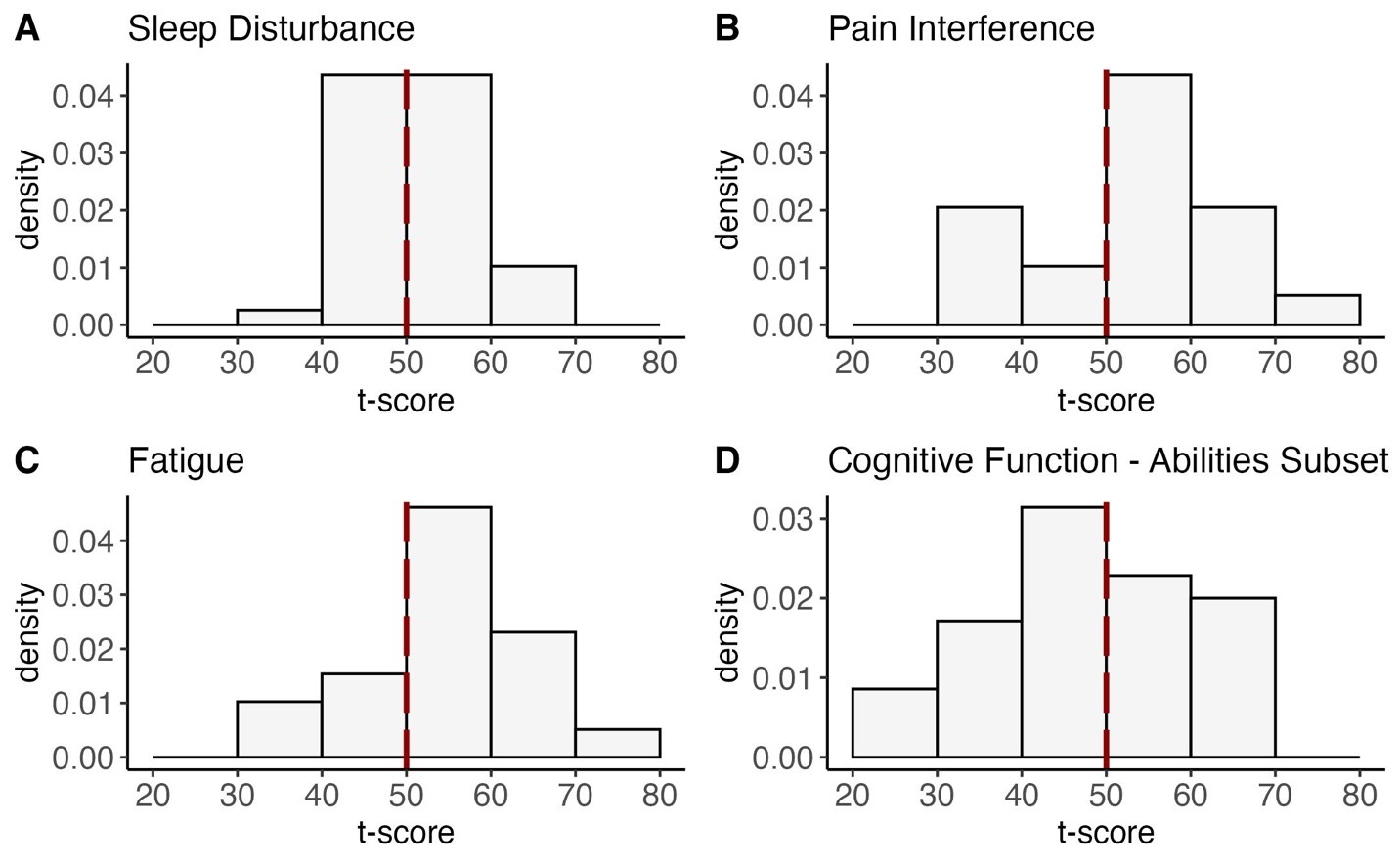
**Figure S4: PROMIS score histograms**. Sleep Disturbance (A), Pain Interference (B), Fatigue (C), and Cognitive Function, Abilities Subset (D) scales are as shown. The reference population is plotted in red as a normal distribution with a mean of 50 and standard deviation of 10.
